# Severity and inpatient mortality of COVID-19 pneumonia from Beta variant infection: a clinical cohort study in Cape Town, South Africa

**DOI:** 10.1101/2021.11.04.21265916

**Authors:** Linda Boloko, Aimee Lifson, Francesca Little, Timothy De Wet, Nectarios Papavarnavas, Gert Marais, Nei-yuan Hsiao, Michael-John Rosslee, Deelan Doolabh, Arash Iranzadeh, Carolyn Williamson, Sipho Dlamini, Marc Mendelson, Ntobeko Ntusi, Robert J. Wilkinson, Hannah Hussey, Mary-Ann Davies, Graeme Meintjes, Sean Wasserman

## Abstract

**Background:** The SARS-CoV-2 Beta variant, associated with immune escape and higher transmissibility, drove a more severe second COVID-19 wave in South Africa. Individual patient level characteristics and outcomes with the Beta variant are not well characterized.

**Methods:** We performed a retrospective cohort study comparing disease severity and inpatient mortality of COVID-19 pneumonia between the first and second wave periods at a referral hospital in Cape Town, South Africa. Beta variant infection was confirmed by genomic sequencing. Outcomes were analyzed with logistic regression and accelerated failure time models.

**Results:** 1,182 patients were included: 571 during the first wave period and 611 from the second wave. Beta variant accounted for 97% of infections in the second wave. There was no difference in crude in-hospital mortality between wave periods (first wave 22.2%, second wave 22.1%; p = 0.9). Time to death was decreased with higher weekly hospital admissions (16%; 95% CI, 8 to 24 for every 50-patient increase), age (18%; 95% CI, 12 to 24 for every 10-year increase) and hypertension (31%; 95% CI, 12 to 46). Corticosteroid use delayed time to death by 2-fold (95% CI, 1.5 to 3.0). Admission during the second wave decreased time to death after adjustment for other predictors, but this did not reach statistical significance (24%; 95% CI, 47 to -2). There was no effect of HIV on survival.

**Conclusions:** There was a trend towards earlier mortality during the second COVID-19 wave driven by the Beta variant, suggesting a possible biological basis. Use of oral prednisone was strongly protective.

**Key points:** In Cape Town, South Africa, the second wave of COVID-19, dominated by the Beta variant, was associated with decreased time to inpatient death after adjustment for age, comorbidities, steroid use, and admission numbers. Use of oral prednisone was strongly protective.

## INTRODUCTION

The causative agent of coronavirus disease 2019 (COVID-19), severe acute respiratory syndrome coronavirus 2 (SARS-CoV-2), has infected hundreds of millions of people across the globe, resulting in over 4.5 million attributed deaths by September 2021 [1]. The first case of COVID-19 in South Africa was identified in March 2020 marking the first wave that peaked in July 2020. At the time of writing, South Africa recorded almost 3 million cases with over 250,000 excess deaths over three epidemic waves [2].

A new variant of concern N501Y.V2, lineage B.1.351 (Beta in the World Health Organization classification), was identified in the Nelson Mandela Bay Metro of the Eastern Cape, South Africa in October 2020 [3]. This variant had eight defining mutations of which three mutations (K417N, N501Y, E484K) were in important sites of the receptor-binding domain (RBD) of the spike protein. By December 2020 the Beta variant was detected in over 90% of viral genomes sequenced in South Africa [4], driving a second wave with higher rates of infections and mortality than those at the peak of the first wave [5].

Mutations associated with the Beta variant result in increased transmissibility because of enhanced human ACE2 (hACE2) receptor binding affinity via the spike protein RBD [6, 7]. In addition to hACE2 receptor binding for entry into human cells, RBD is the target of SARS-CoV-2 neutralizing antibodies during natural infection [8]. *In vitro* studies using convalescent plasma from people infected with earlier variants during the first wave demonstrated a reduction in the neutralization capacity against the Beta variant [9]. Both immune escape and higher transmissibility have been proposed as explanations for a more severe second wave in South Africa.

Epidemiological data from three South African districts suggested higher in-hospital mortality in the second wave compared to the first wave [5]. However, studies comparing individual level characteristics and outcomes by variant are lacking and are needed to inform patient management and public health planning. We undertook a retrospective cohort study to characterize clinical phenotype and compare mortality in patients admitted to hospital during the first and second waves, predominated by ancestral variants and the Beta variant, respectively.

## METHODS

### Study population

We identified patients 18 years or older admitted to Groote Schuur Hospital, a large referral hospital in Cape Town, South Africa, with COVID-19 pneumonia, defined as respiratory illness confirmed by SARS-CoV-2 by reverse transcriptase polymerase chain reaction (RT-PCR) on a respiratory sample. Patients were included from two distinct periods: the first wave, 26 March to 10 July 2020; and the second wave, 15 November 2020 to 15 January 2021. These dates were selected to represent a complete time profile of each wave (different phases may influence hospital outcomes) and to coincide with the period at which point the Beta variant was the dominant variant in circulation during the second wave. Screening for potentially eligible cases was done on randomly selected medical records generated from lists of consecutive hospital COVID-19 admissions during each study period. The management of COVID-19 is standardized in our hospital [10]. Corticosteroid use, mainly prednisone 40 mg daily, was introduced to management protocols for all patients with COVID-19 pneumonia on 16 June 2020 after publication of the RECOVERY trial results [11]. Although some patients initially received intravenous dexamethasone, the vast majority were treated with oral prednisone as oral dexamethasone is not available in South Africa. Low molecular weight heparin was provided to all patients, either at intensified (1 mg/kg daily or twice daily) or prophylactic doses (0.5 mg/kg daily), in line with a hospital guideline [10]. No patients received remdesivir, IL-6 inhibitors, or monoclonal antibodies. Allocation of intensive care unit (ICU) beds and high flow oxygen was based on a triage scoring tool developed by the provincial Department of Health [12] and resource availability. The national vaccination program had not yet started at the end of the data collection period.

### Data sources

We collected baseline characteristics including demographics, symptom type and duration, and comorbidities. Anthropometry was based on clinical observation or direct measurements in those able to stand; because of poorly documented weight measurements we recorded weight in three categories as indicated in the clinical notes: underweight, normal weight, or overweight. Routine bedside and laboratory investigations of potential prognostic importance were captured. We also extracted data on medication, oxygen requirements, including use of high flow nasal oxygen (HFNO), and ICU admission. Discharge from hospital or death during the index admission was ascertained directly from medical records or via the electronic hospital clinical management system. Data was captured from medical records directly onto electronic case report forms designed specifically for this study and exported to statistical analysis software. We obtained total COVID-19 admissions numbers from the Provincial Health Data Centre, a database repository used by the Western Cape Provincial Department of Health that integrates electronic data from multiple data sources including laboratory results and clinical episodes at provincial facilities and hospitals [13].

### SARS-CoV-2 testing and sequencing

All SARS-CoV-2 RT-PCR testing at Groote Schuur and its public sector referral hospitals was done by the on-site National Health Laboratory Services (NHLS) diagnostic laboratory. We also included patients initially tested in private laboratories prior to presentation to Groote Schuur. The Allplex™ 2019-nCoV assay (Seegene, South Korea) kit was almost exclusively used for testing in the public sector. This kit targets the E gene, RdRp gene and the N gene of SARS-CoV-2. A cycle threshold (Ct) value ≤40 in any target gene was considered positive. We selected the N gene for Ct value analysis as no mutations associated with altered primer binding efficiency were prevalent in the SARS-CoV-2 lineages circulating at the time of the study.

To confirm presence of the Beta variant, we performed whole genome sequencing of second wave SARS-CoV-2 samples that were available from storage at the University of Cape Town and NHLS Division of Medical Virology. Genetic sequencing was not done on first wave samples as there were no reported variants of concern (VOC) until October 2020 and our study period ended in July 2020 for the first wave [4]. The sequencing method is described the supplementary material (Supplementary text 1)

### Statistical analysis

The primary outcome measure was the proportion of patients experiencing in-hospital mortality, with wave period as the main exposure variable. A power calculation was done based on data collected during the first wave, where in-hospital mortality was 22% among a sample of 571 patients. Outcomes data from similar number of patients would provide more than 80% power to detect 1.5-fold higher odds of mortality (7% absolute increase) in a cohort admitted during the second wave and infected with the Beta variant.

Baseline characteristics are presented as proportions for categorical variables and medians with interquartile range for continuous variables and compared by wave using rank sum test and Fisher exact test and rank sum test, respectively. We used two approaches to evaluate whether admission during the second wave, as a proxy for infection with the Beta variant, was associated with inpatient mortality. First, we did multivariable logistic regression to compare overall mortality over the observed periods, adjusting for age, sex, comorbidities, and steroid use. We also adjusted for total number of COVID-19 hospital admissions over each period as a measure of health system pressure which may influence outcome. Second, we performed survival analysis to incorporate time to death and censoring at day 28 after admission. We fitted a gamma regression accelerated failure time model to analyze time to the primary outcome, adjusted for the same potential confounders as included in the logistic regression models. Analysis was performed in Stata 12 and R version 4.0.1. This study was approved by the University of Cape Town Human Research Ethics Committee (HREC 285/2020).

## RESULTS

### Clinical and viral characteristics

1,182 patients were included: 571 during the first wave period and 611 from the second wave. Most had severe respiratory illness, with median partial pressure of arterial oxygen (PaO2) 8.3 kPa (IQR 6.7 - 10.8) measured on room air or supplemental oxygen, and evidence of systemic inflammation with elevated median C-reactive protein (CRP) and D-dimer values, the latter being lower among second wave patients (Table 1). Notable differences in the second wave cohort with respect to the first wave included slightly older age, and higher prevalence of diabetes mellitus (45% vs 35%), hypertension (55% vs 49%) and being overweight (46% vs 40%). 117 participants were HIV-positive, with a higher proportion among those admitted in the first wave (15% vs 8% in the second wave), although there was substantial missing data on HIV status (n = 198) in the second wave. Median time to hospital presentation from symptom onset was shorter in the first wave than in the second wave (5 days, IQR 3-7, vs. 6 days, IQR 4-8). Cough and dyspnea were more frequently reported in the second wave, but other presenting symptoms were similar (figure 1 & figure S1).

**Table 1:** Baseline characteristics.

|  | **First wave**<br>**n=571** | **Second wave**<br>**n=611** | **Overall**<br>**N=1182** | **P value** |
| --- | --- | --- | --- | --- |
| **Age, years (median, IQR)** | 53 (41-65) | 58 ( 47-68) | 56 (45-67) | 0.001 |
| **Male sex (n, %)** | 241 (42%) | 281 (46%) | 522 (44%) | 0.191 |
| **Overweight (n, %)** | 228 (40%) | 284 (46%) | 512 (43%) | 0.023 |
| **HIV<sup>a</sup> positive (n, %)** | 83 (15%) | 34/413 (8%) | 117/984 (12%) | 0.003 |
| **Diabetes mellitus(n, %)** | 199 (35%) | 273 (45%) | 472 (40%) | 0.001 |
| **Hypertension** | 279 (49%) | 335 (55%) | 614 (52%) | 0.042 |
| **Past tuberculosis (n, %)** | 51 (9%) | 23 (4%) | 74 (6%) | 0.001 |
| **Active tuberculosis (n, %)** | 17 (3%) | 12 (2%) | 29 (2%) | 0.260 |
| **In HIV positive patients:** |  |  |  |  |
| **- On ART<sup>b</sup>** | 69 (83%) | 26 (76%) | 95 (81%) | 0.402 |
| **-CD4 cells/mm3** | n=73 | n=19 | n=92 |  |
| **median (IQR)** | 295 (129-471) | 192 (89-227) | 237 (111-452.5) | 0.071 |
| **- log10(HIV VL)** | n=21 | n=10 | n=31 |  |
| **median (IQR)** | 2.3 (1.7-3,2) | 4.1 (2.1-5.7) | 2.4 (1.7-5.7) | 0.069 |
| **Symptoms (n, %)** |  |  |  |  |
| **Cough** | 380 (67%) | 458 (75%) | 838 (71%) | 0.001 |
| **Dyspnoea** | 421 (74%) | 549 (90%) | 970 (82%) | <0.001 |
| **Sore throat** | 120 (21%) | 109 (18%) | 229 (19%) | 0.167 |
| **Myalgia** | 143 (25%) | 149 (24%) | 292 (25%) | 0.793 |
| **Fever** | 264 (46%) | 248 (41%) | 512 (43%) | 0.050 |
| **Oxygen saturation (%)** | n=553 | n=523 | n=1076 |  |
| **median (IQR)** | 93 (88-96) | 88 (82-92) | 90 (84-95) | <0.001 |
| **PaO<sub>2</sub> (kPa)** | n=321 | n=539 | n=860 |  |
| **Median (IQR)** | 8 (6.9-10.7) | 8.5 (6.9-10.7) | 8.3 (6.7-10.8) | 0.041 |
| **White cell count** |  |  |  |  |
| – **Neutrophils (10<sup>9</sup>/L)** | n=416 | n=390 | n=806 |  |
| **median (IQR)** | 5.8 (4.1-8.7) | 6.1 (4.3-8.7) | 5.9 (4.2-8.7) | 0.949 |
| – **Lymphocytes (10<sup>9</sup>/L)** | n=419 | n=390 | n=809 |  |
| **median (IQR)** | 1.4 (0.97-2) | 1.2 (0.79-1.8) | 1.3 (0.9-1.9) | 0.001 |
| **CRP, mmol/l** | n=351 | n=354 | n=705 |  |
| **median (IQR)** | 93 (43-182) | 100 (51-149) | 97 (47-161) | 0.662 |
| **D-dimer (mg/L)** | n=310 | n=488 | n=798 |  |
| **Median (IQR)** | 0.7 (0.4-1.4) | 0.5 (0.3-0.9) | 0.6 (0.3-1.0) | 0.002 |

<sup>a</sup>human immunodeficiency virus, <sup>b</sup>antiretroviral therapy

**Figure 1.**
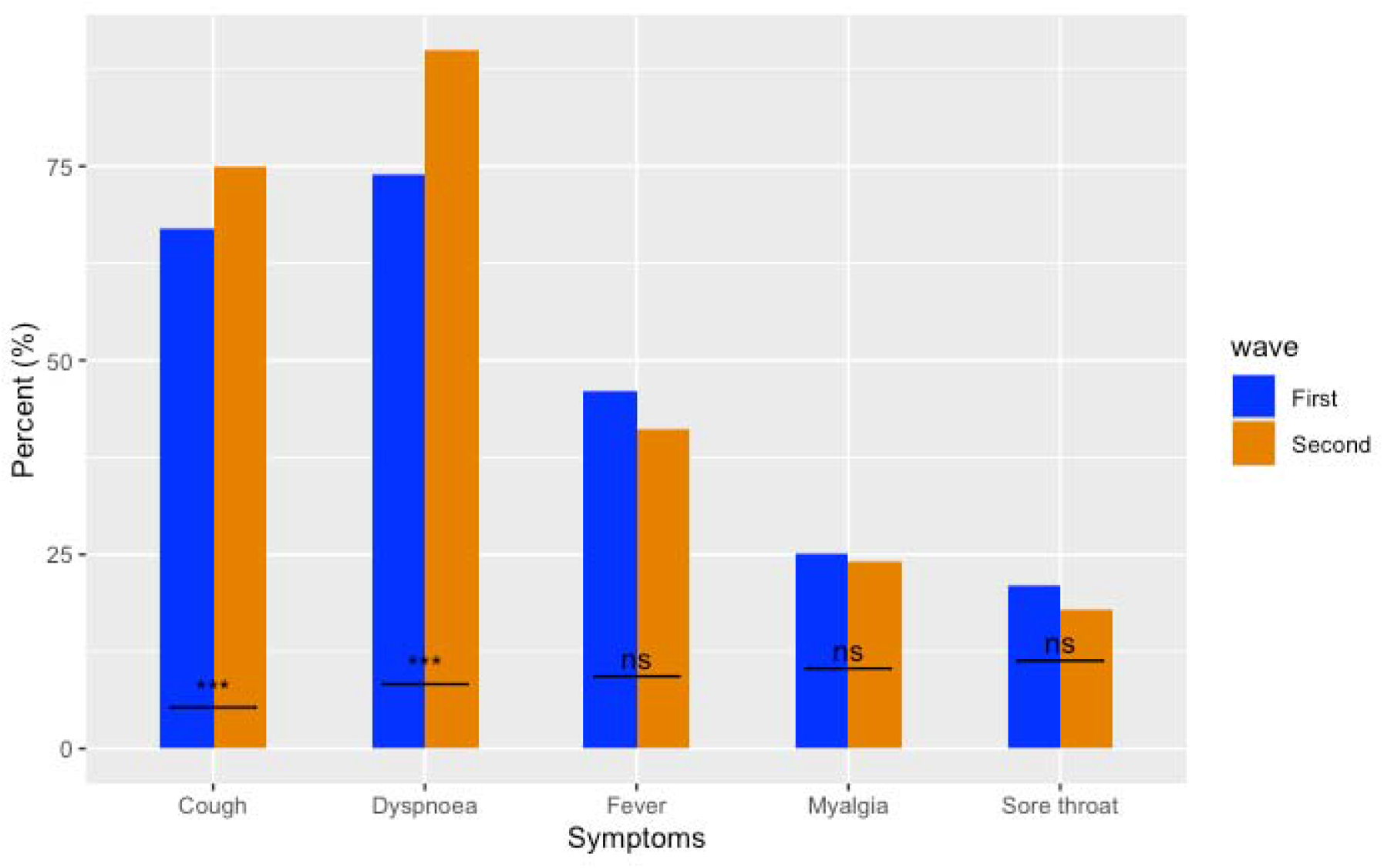
Proportions of symptoms presented by first and second wave. *** p<0.001, ns: not significant.

The second wave had a higher peak and more rapid increase in hospital admissions per week than the first wave (figure S2). Sequencing data was available for 117 (19.1%) samples collected from patients during the second wave; 113 (97%) were confirmed to be Beta variant (figure 2). There was no difference in Ct values by wave (median Ct first wave 31.1, IQR 26.5 to 34.4 vs. median Ct second wave 30.7, IQR 27.4 to 34.1; p = 0.6), but samples from patients who died had lower N gene Ct values compared to those who survived (median Ct 28.7, IQR 26 to 32.9 vs median Ct 31.4, IQR 27.7 to 34.5; p <0.001) (figure S3).

**Figure 2.**
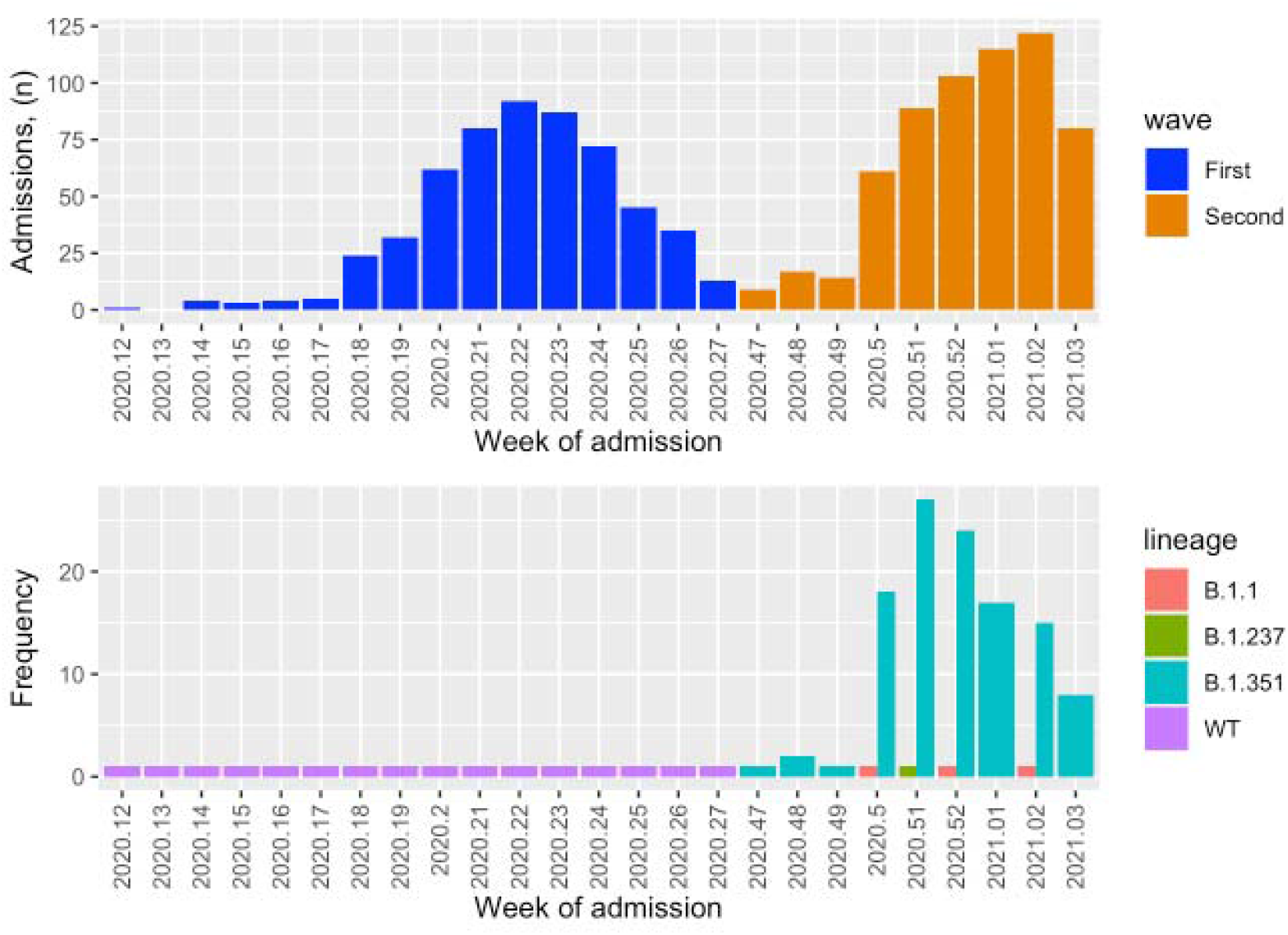
COVID-19 admissions to hospital and viral sequencing over the study periods. *Top panel*: Total number of study population admissions each week of the year studied comparing the first and the second wave. *Bottom panel*: Corresponding number of confirmed variants for participants sequenced in the second wave. WT: wild type

### Management and outcomes of patients hospitalized with COVID-19

In the second wave, more patients were managed with HFNO (15.4% vs 8.6%), corticosteroids (93% vs 14%), and intensified anticoagulation (94% vs 63%) compared with the first wave. More patients were admitted to ICU and received invasive mechanical ventilation in the first wave (9% vs 5.9% in the second wave). Duration of hospital stay was similar in those surviving to hospital discharge (Table 2).

**Table 2.** In-hospital management and outcomes.

|  | **First Wave**<br>**(n=571)** | **Second Wave**<br>**(n=611)** | **Total**<br>**(N=1182)** | **p value** |
| --- | --- | --- | --- | --- |
| **HFNO<sup>a</sup>** | 49 (8.6%) | 94 (15.4%) | 143 (12.1%) | < 0.001 |
| **ICU<sup>b</sup> admission** | 54 (9%) | 36 (5.9%) | 71 (6.0%) | 0.021 |
| **Corticosteroid use** | 78 (13.7%) | 566 (92.6%) | 644 (54.5%) | < 0.001 |
| **Intensified**<br>**anticoagulation** | 358 (62.7%) | 571<br>(93.5%) | 929 (78.6%) | < 0.001 |
| **Time to discharge, days**<br>**(median, IQR)** | 7 (4-12) | 6 (4-10) | 6 (4-11) | 0.5 |
| **Time to death, days**<br>**(median, IQR)** | 6 (3-9) | 6 (3-12) | 6 (3-10) | 0.4 |
| **Deaths** | 127 (22.2%) | 135 (22.1%) | 262 (22.2%) | 0.9 |

<sup>a</sup>High flow nasal oxygen, <sup>b</sup>intensive care unit

### In-hospital mortality

Crude in-hospital mortality was similar between wave periods (first wave 22.2%, second wave 22.1%) (Table 2). However, there was a stronger relationship between total weekly COVID-19 hospital admissions and total weekly COVID-19 deaths in the second wave compared to the first wave (first wave rho = 0.55; p = 0.03 vs second wave rho = 0.84; p = 0.004) (figure S4). Patients who died were older, more likely to be male, had more frequent hypertension and kidney disease, and markers of disease severity, including poorer oxygenation, higher neutrophil count, CRP, and D-dimer concentrations (Table S1). Admission during the second wave was not associated with increased risk of inpatient mortality after adjustment for steroid use, total admissions, age, and comorbidities on multivariable logistic regression (aOR 0.97; 95% CI 0.6 to 1.7). Steroid use was associated with much lower odds of inpatient mortality (aOR 0.48; 95% CI, 0.3 to 0.8); total number of admissions had the opposite effect, with 1.2-fold (95% CI, 1.1 to 1.4) increased odds of death for every 50-person increase in weekly hospital admissions (Table 3).

**Table 3:** Multivariable logistic regression model.

|  | **Adjusted odds ratio** | **95% confidence interval** | **P value** |
| --- | --- | --- | --- |
| **Second wave period** | 0.97 | 0.55 to 1.7 | 0.9 |
| **COVID-19 admission (50-person increase per week)** | 1.2 | 1.1 to 1.4 | 0.009 |
| **Age (every 10 year increase)** | 1.5 | 1.3 to 1.7 | <0.001 |
| **Sex** |  |  |  |
| - Female | 1 | 1 | 1 |
| - Male | 1.4 | 1.1 to 1.94 | 0.02 |
| **No hypertension and diabetes** | 1 | 1 | 1 |
| **Hypertension** | 1.8 | 1.1 to 2.7 | 0.01 |
| **Diabetes** | 0.76 | 0.43 to 1.4 | 0.3 |
| **Both diabetes and hypertension** | 1.3 | 0.87 to 2.0 | 0.2 |
| **Chronic kidney disease** | 2.0 | 1.2 to 3.1 | 0.004 |
| **ICU<sup>a</sup> admission** | 5.7 | 3.3 to 9.6 | <0.001 |
| **HFNO<sup>b</sup>** | 4.2 | 2.6 to 6.7 | <0.001 |
| **Corticosteroid use** | 0.4 | 0.28 to 0.84 | 0.01 |

<sup>a</sup>intensive care unit, <sup>b</sup>high flow nasal oxygenation

### Time to in-hospital mortality

An adjusted AFT model estimated that time to death was decreased by 16% (95% CI, 8 to 24) for every 50-person increase in weekly COVID-19 hospital admissions. Admission during the second wave also decreased time to death, but this did not reach statistical significance (24%; 95% CI, 47 to -2). Known poor prognostic factors, including age (18%; 95% CI, 12 to 24 for every 10-year increase), hypertension (31%; 95% CI, 12 to 46), and chronic kidney disease (28%; 95% CI, 6 to 44), also independently decreased time to death. Use of steroids delayed time to death by 2-fold (95% CI, 1.7 to 3) (Figure 3 & Table S2). There was no effect of HIV on survival time (including on sensitivity analysis when missing values were imputed as negative). In the final model, cumulative hazard of inpatient death for an average patient in the second wave was higher than in the first wave, with a large and significant protective effect from steroid use (Figure 4).

**Figure 3.**
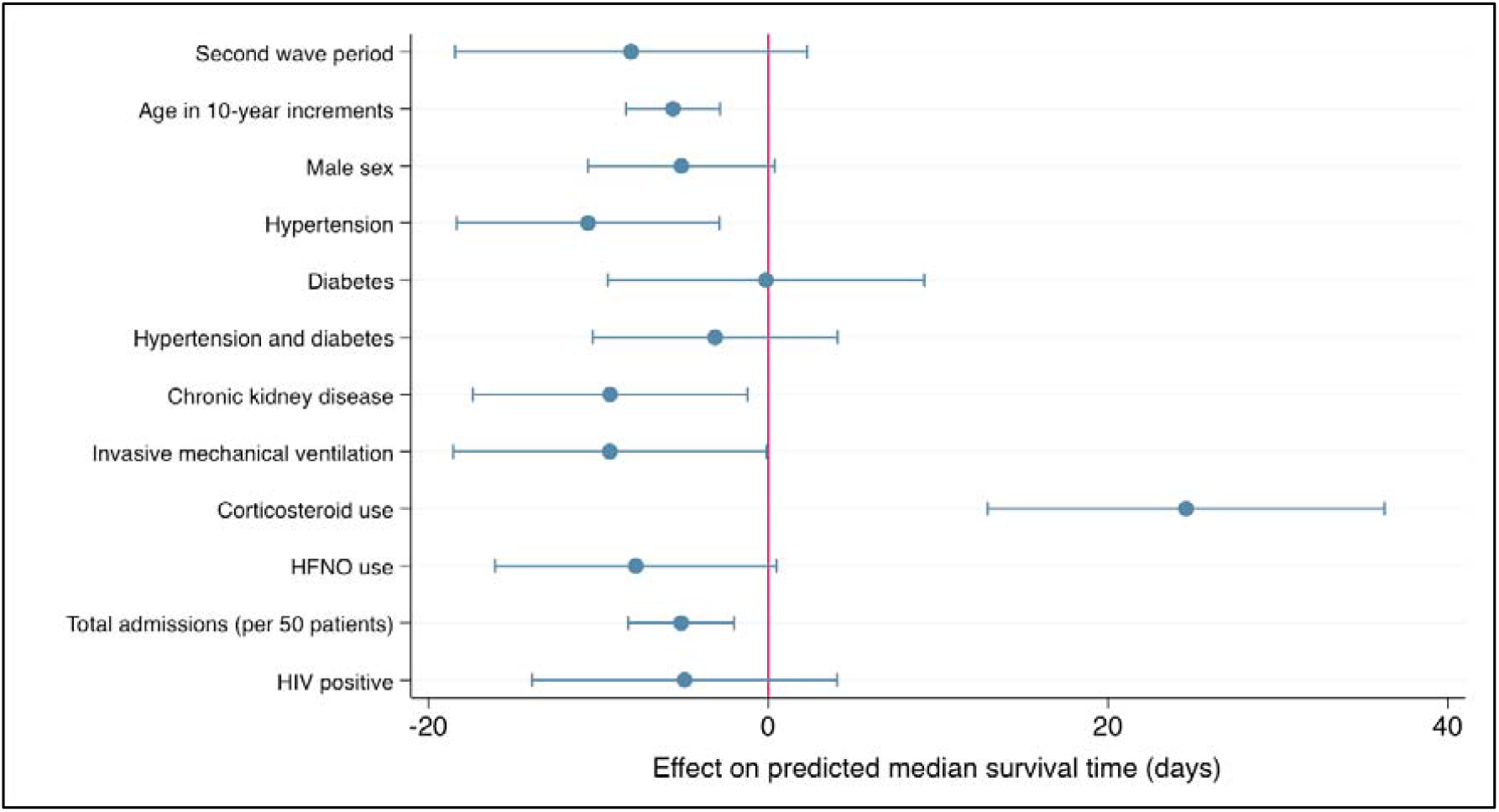
Effect of covariates on in-hospital mortality from accelerated failure time. model Circles are point estimates, bars are 95% confidence intervals, of average marginal effects. Vertical red line indicates no effect. HFNO, high flow nasal oxygen. HIV data are imputed for 198 patients (missing coded as negative).

**Figure 4.**
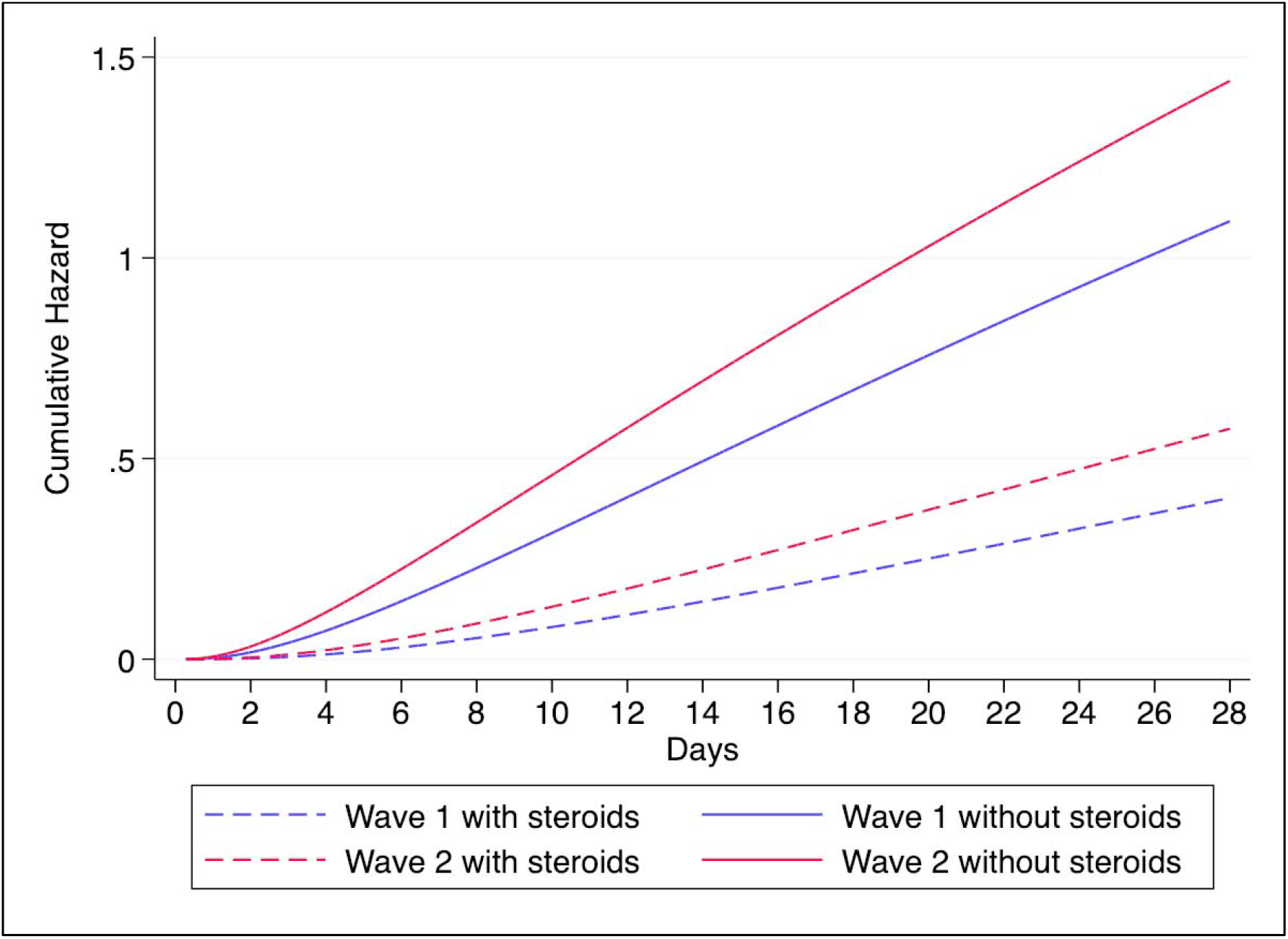
Cumulative hazard function for mortality. Cumulative hazard rate for mortality over time is higher for the second wave compared to the first wave at mean values of baseline covariates.

## DISCUSSION

In this retrospective cohort study of hospitalized patients with COVID-19 in Cape Town, South Africa, clinical severity and inpatient mortality during the second wave, dominated by the Beta variant, was similar to the first wave period. However, there was a signal that time to death was faster in the second wave cohort after accounting for steroid use, which had a large protective effect, and for total hospital admissions, which was associated with a higher risk for mortality.

In contrast to our patient-level data, population studies have reported clear increases in mortality with Beta and other VOC. A large population-based analysis in South Africa showed that the second wave was associated with a 31% residual increased risk of in-hospital mortality after adjustment for important epidemiological factors [5]. A significantly higher risk for severe illness and mortality was also seen with Beta variant infection relative to Alpha (B.1.1.7) in the Qatari national COVID-19 database, but residual confounding was a concern [14]. In the United Kingdom a series of studies suggested that the Alpha variant conferred between 55% and 73% higher risk of mortality compared to ancestral variants [15-17]. However, this was not seen in smaller hospitalized cohorts with more granular clinical data [18, 19], and importantly, less confounding related to resource limitations in these higher income settings. Disentangling an isolated effect of VOC from dynamic factors that strongly influence clinical outcomes, such as health system strain, treatment practices, vaccination, and population behaviors, is challenging [20].

Laboratory data have demonstrated that variants with mutations in the receptor-binding domain including Alpha, Beta, and Gamma likely have increased transmissibility and reduced sensitivity to neutralization by natural and vaccine-induced antibodies [6, 9, 21, 22]. These characteristics have resulted in higher infection peaks, and increased hospitalization and mortality associated with these variants, including in South Africa [2, 23, 24]. High viral loads may further predict the probability of progression of disease during hospitalization [25], as shown in our study those who died had higher viral loads than survivors. Although lower RT-PCR amplification cycles have been reported with VOC, including the Beta variant [26], a causal relationship between Ct values, VOC, and mortality is not yet established. Dynamic measures of Ct values provide better insights into VOC biology. We did not have longitudinal viral testing data and similar Ct values between wave periods observed in our study may have been influenced by time course of illness or sampling differences.

In line with international experiences [27, 28], we found a strong relationship between hospital deaths and total weekly admissions, reflecting the influence of health system pressure on inpatient outcomes, particularly in hospitals with limited resources such as ours [29, 30]. HFNO has been shown to reduce the need for mechanical ventilation and ICU admission [31]. The higher proportion managed with HFNO in the second wave reflects increased use of this intervention to mitigate the need for mechanical ventilation in the context of severe ICU bed constraints.

Use of corticosteroids is associated with survival benefit in hospitalized patients with COVID-19 [11, 32]. High quality evidence only exists for dexamethasone; international guidelines recommend use of other corticosteroids based on dose equivalencies. Methylprednisolone, which has similar potency to prednisone, has been studied mainly in small case series or retrospective cohorts at variable doses; a recent systematic review only found one published case series of prednisone in COVID-19 (n = 6). Our study showed that prednisone use reduced risk of mortality by 46% and 2-fold decelerated time to death, providing support for use where dexamethasone is unavailable.

Previous studies have shown diabetes mellitus to be an independent predictor of mortality in patients with COVID-19, with increasing risk in poorly controlled disease [33, 34]. We did not find this effect. A potential explanation is that patients with diabetes mellitus may have received intensified care including improved glucose control from protocols and interventions developed in our institution and province, respectively [10, 35]. Well-controlled in-hospital glucose concentrations during COVID-19 associates with reduced risk of mortality [36]. However, an unmeasured confounder that may also explain this finding is the widespread use of metformin in our population which has been associated with lower mortality in diabetic patients with COVID-19 prescribed this agent prior to admission compared to other oral diabetic agents [37, 38]. We did not ascertain whether patients had newly diagnosed diabetes mellitus, which may have a lower risk of mortality, diluting the overall effect.

Limitations of our study include retrospective medical record review which may have led to selection bias from inconsistent record retrieval, data quality issues including inaccurate weight measurements, and missing data. We did not adjust for the effect of non-COVID-19 admissions which may have resulted in further pressures on hospital systems. Increased corticosteroid use in the second wave may have masked a higher intrinsic risk of mortality from the Beta variant despite adjusting for it in our models. Our hospital is a referral centre and may not reflect the general population, limiting generalizability. And finally, despite inclusion of important patient-level predictors in our models, there may be unmeasured residual confounding. Our study has several strengths that support the conclusions. First, there were many events allowing for more precise assessment of mortality outcomes and providing sufficient power to adjust for multiple confounders. Our patient-level data provides more granular information with advantages over community-based studies. Second, we did not include vaccinated patients in our cohort which would have influenced severity outcomes. Third, we performed viral sequencing in the second wave to validate the use of the wave period as a reliable proxy for Beta variant infection.

## Conclusions

In line with other experience, we showed that hospital admission pressure is an important factor in outcomes during COVID-19 epidemic waves. After adjusting for increased hospital admissions and other important patient-level covariates, there was a trend towards accelerated mortality during the second COVID-19 wave driven by the Beta variant, suggesting a possible biological effect. Use of oral prednisone was strongly protective and should be considered as an alternative to intravenous corticosteroids or dexamethasone for COVID-19 pneumonia in low resource settings. Current vaccines remain protective against severe outcomes with Beta variant infection [39] and expansion of the vaccination program is a priority.

## Data Availability

All data produced in the present study are available upon reasonable request to the authors

## ACKNOWLEDGEMENTS

We acknowledge and thank all frontline staff at Groote Schuur Hospital who contributed to the clinical service providing outstanding care to our patients.

