## Supplemental table S1 and figure S1-S4 for "Severity and inpatient mortality of COVID-19 pneumonia from Beta variant infection: a clinical cohort study in Cape Town, South Africa"

Supplementary material

**Supplementary text 1**: Sequencing methods

**Figure S1**: Total hospital weekly admissions during the first and second wave

**Figure S2**: Comparison of Ct values by mortality

**Figure S3**: Presenting symptoms

**Figure S4**: Scatterplot of relationship between hospital admission and deaths by wave

**Table S1**: Clinical features of survivors and non-survivors

**Supplementary text 1: Sequencing method**

Briefly, RNA was extracted from nasopharyngeal swabs of qPCR-confirmed COVID-19 patients using automated methods. Complementary DNA was synthesized using the Superscript IV reverse transcriptase (Life Technologies, Carlsbad, CA) and random hexamer primers. SARS-CoV-2 whole genome amplification was performed using the ARTIC V3 protocol [1].

In summary, multiplex PCR was performed using primers designed on Primal Scheme (<http://primal.zibraproject.org/>) to generate 400 base pair (bp) amplicons with a 70 bp overlap covering the SARS-CoV-2 genome. PCR products were purified using AMPure XP magnetic beads (Beckman Coulter, CA) and quantified using the Qubit dsDNA High Sensitivity assay on the Qubit 3.0 instrument (Life Technologies Carlsbad, CA). The Illumina® DNA Prep kit with Nextera DNA CD Indexes (96 indexes) was used to prepare indexed paired-end libraries of genomic DNA. Sequencing libraries were normalized to 4 nM, pooled and denatured with 0.2 N sodium hydroxide. Libraries were sequenced on the Illumina MiSeq instrument (Illumina, San Diego, CA, USA). Paired-end fastq reads were assembled using Genome Detective 1.132 (<https://www.genomedetective.com>) and the Coronavirus Typing Tool. The assembly obtained from Genome Detective were submitted to the Nextclade webpage (<https://clades.nextstrain.org>) for the initial quality assessment. Genomes were polished by aligning mapped reads to the references and filtering out low-quality mutations using bcftools 1.7-2 mpileup method. Mutations were confirmed visually with bam files using Geneious software (Biomatters Ltd, New Zealand). Any assembled Genomes with missing sites greater than 10% of the reference genome size were removed from the dataset (> 3000 base pairs). To assign the sequenced samples to their lineage and clade, we used the dynamic lineage classification method proposed by Rambault et al. via the Phylogenetic Assignment of named Global Outbreak LINeages (PANGOLIN) software suite (https://github.com/hCoV2019/pangolin) 4 , and Nextclade5 , respectively.

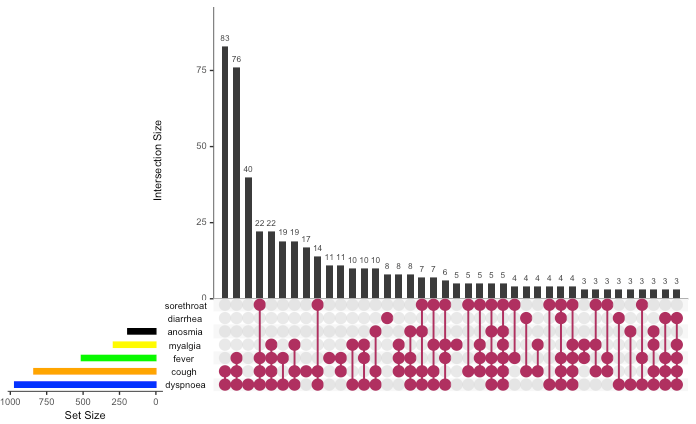

**Figure S1:** Total set size (number of patients with cough, dyspnoea, fever, myalgia, sore throat, diarrhea and anosmia) shown by horizontal coloured bars. Intersections of these sets are indicated by the connected purple dots; number of patients in each of the possible intersections are shown with vertical bars.

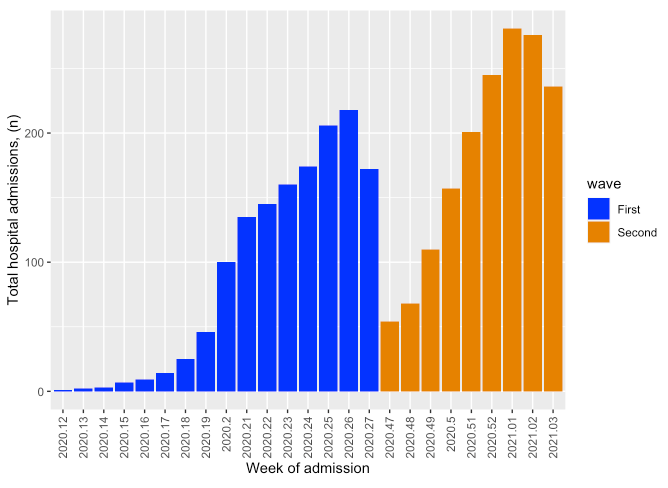

**Figure S2**: Comparison of first wave and second wave total hospital weekly admissions in the periods studied.

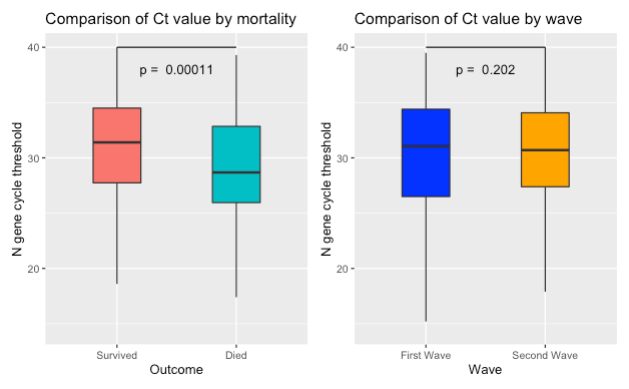

**Figure S3**: Comparison of viral load by proxy using Ct values. *Left panel*: Lower Ct values in those who died compared to survivors. *Right panel*: No difference in Ct values between the waves.

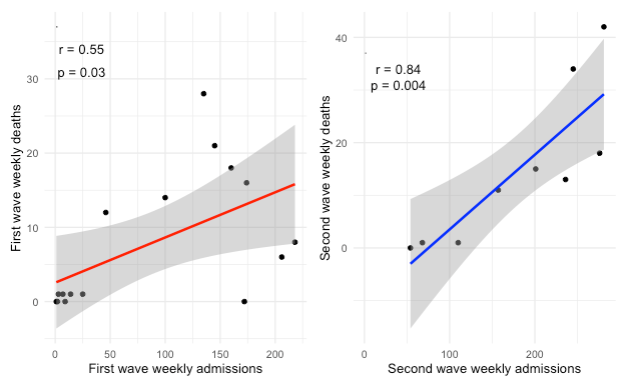

**Figure S4:** Correlation and linear regression of hospital weekly admissions and weekly deaths by wave of admission..

**Table S1**: Clinical characteristics of survivors vs non-survivors

|  | **Died (n = 262)** | **Survived (n= 920)** | **Total (N=1182)** | **p value** |
| --- | --- | --- | --- | --- |
| **Age, years**  **Median (IQR)** | 62 (52-71) | 54 (43-65) | 56 (45-67) | < 0.001 |
| **Male sex** | 134 (51.1%) | 388 (42.2%) | 522 (44.2%) | 0.01 |
| **Hypertension** | 170 (64.9%) | 444(48.3%) | 614 (51.9%) | < 0.001 |
| **Diabetes mellitus** | 113 (43.1%) | 359 (39%) | 472 (39.9%) | 0.2 |
| **Chronic kidney disease** | 42 (16%) | 67 (7.3%) | 109 (9.2%) | <0.001 |
| **HIV** | 27 (12.1%) | 90 (11.8%) | 117 (11.9%) | 0.9 |
| **Past TB** | 18 (6.9%) | 56 (6.1%) | 74 (6.3%) | 0.6 |
| **Saturation** | 88% (83-92) | 94 (90-97) | 93 (88-96) | <0.001 |
| **pO2** | 7.14 (5.8-9) | 8.6 (7-11.5) | 8.3 (6.7-10.8) | <0.001 |
| **White cell count**   - **Neutrophils** - **Lymphocytes** - **Platelets**   **Creatinine**  **CRP**  **D-dimer (<0.25)** | 9.0 (6.5-12.4)  7.8 (5.5-11.8)  1.3 (0.9-1.8)  228 (176-302)  106 (81-155)  127 (70-239)  0.68 (0.42-1.8) | 7.6 (5.8-10.0)  5.5 (3.9-8.1)  1.4 (1.0-2.0)  247 (198-14)  81 (64-105)  89 (42-149)  0.55 (0.32-0.88) | 7.9 (5.9-10.5)  5.8 (4.1-8.7)  1.4 (1.0-2.0)  243 (192-312)  86 (66-112)  97 (47-161)  0.6 (0.34-1) | <0.001  <0.001  0.05  0.007  <0.001  <0.001  <0.001 |

**Table S2**: Adjusted accelerated failure time model estimates

|  | **Time ratio** | **95% CI** | **P value** |
| --- | --- | --- | --- |
| **Weekly hospital admission (every 50-patients increase)** | 0.84 | 0.76 to 0.92 | <0.001 |
| **Second wave** | 0.76 | 0.53 to 1.1 | 0.1 |
| **Age (every 10 year increase)** | 0.82 | 0.76 to 0.89 | <0.001 |
| **Male sex** | 0.84 | 0.7 to 1 | 0.06 |
| **Hypertension without diabetes mellitus** | 0.69 | 0.54 to 0.89 | 0.004 |
| **Diabetes mellitus without hypertension** | 1 | 0.72 to 1.3 | 1 |
| **Both diabetes and hypertension** | 0.9 | 0.70 to 1.1 | 0.4 |
| **Chronic kidney disease** | 0.72 | 0.56 to 0.94 | 0.02 |
| **ICU admission** | 0.72 | 0.53 to 0.98 | 0.04 |
| **HFNO^a^** | 0.76 | 0.58 to 0.99 | 0.05 |
| **Corticosteroid use** | 2.3 | 1.7 to 3.3 | <0.001 |
| **HIV^b^** | 0.84 | 0.62 to 1.1 | 0.3 |

^a^high flow nasal oxygenation, ^b^HIV: human immunodeficiency virus
